# Molecular diagnostic yield of genome sequencing versus targeted gene panel testing in racially and ethnically diverse pediatric patients

**DOI:** 10.1101/2023.03.18.23286992

**Authors:** Noura S. Abul-Husn, Priya N. Marathe, Nicole R. Kelly, Katherine E. Bonini, Monisha Sebastin, Jacqueline A. Odgis, Avinash Abhyankar, Kaitlyn Brown, Miranda Di Biase, Katie M. Gallagher, Saurav Guha, Nicolette Ioele, Volkan Okur, Michelle A. Ramos, Jessica E. Rodriguez, Atteeq U. Rehman, Amanda Thomas-Wilson, Lisa Edelmann, Randi E. Zinberg, George A. Diaz, John M. Greally, Vaidehi Jobanputra, Sabrina A. Suckiel, Carol R. Horowitz, Melissa P. Wasserstein, Eimear E. Kenny, Bruce D. Gelb

## Abstract

**Purpose:** Adoption of genome sequencing (GS) as a first-line test requires evaluation of its diagnostic yield. We evaluated the GS and targeted gene panel (TGP) testing in diverse pediatric patients (probands) with suspected genetic conditions.

**Methods:** Probands with neurologic, cardiac, or immunologic conditions were offered GS and TGP testing. Diagnostic yield was compared using a fully paired study design.

**Results:** 645 probands (median age 9 years) underwent genetic testing, and 113 (17.5%) received a molecular diagnosis. Among 642 probands with both GS and TGP testing, GS yielded 106 (16.5%) and TGPs yielded 52 (8.1%) diagnoses (*P* < .001). Yield was greater for GS *vs*. TGPs in Hispanic/Latino(a) (17.2% *vs*. 9.5%, *P* < .001) and White/European American (19.8% *vs*. 7.9%, *P* < .001), but not in Black/African American (11.5% *vs*. 7.7%, *P* = .22) population groups by self-report. A higher rate of inconclusive results was seen in the Black/African American (63.8%) *vs*. White/European American (47.6%; *P* = .01) population group. Most causal copy number variants (17 of 19) and mosaic variants (6 of 8) were detected only by GS.

**Conclusion:** GS may yield up to twice as many diagnoses in pediatric patients compared to TGP testing, but not yet across all population groups.

## INTRODUCTION

Genetic testing plays an important role in pediatric medicine and is increasingly being used by geneticists and non-genetics specialists to diagnose a variety of conditions. Paradigms for diagnostic genetic testing are variable, can include several testing modalities, and continue to evolve with advancing genomic sequencing technologies.^1^ Contemporary clinical testing often includes the use of targeted gene panels (TGPs), which consist of sets of genes that have been causally implicated in a particular phenotype. In order to select among commercially available TGPs, the ordering physician must have some suspicion of the genes of interest based on the clinical phenotype. The uptake of unbiased, genome-wide approaches using clinical exome (ES) or genome (GS) sequencing has steadily increased over the last decade,^2,3^ with these approaches avoiding the need for clinical suspicion of a particular disease or set of genes.

The broad scope of GS offers the greatest potential to increase the diagnostic yield for individuals with suspected genetic conditions.^4–9^ GS allows for the analysis of single nucleotide variants (SNVs) and insertion/deletions (indels) in all genes with more uniform sequencing coverage than ES. GS further enables the detection of variants in intronic and other non-coding regions of the genome, copy number variants (CNVs), mitochondrial variants, and other types of variants that can be missed by other test modalities.^10,11^ Additional benefits of GS include reducing the time to achieve a diagnosis by avoiding the need for multiple sequential tests and the possibility to reanalyze GS data as new knowledge is gained with respect to gene-disease associations and variant level interpretation.^12^ However, the clinical utility and cost-effectiveness of GS as a first-line diagnostic test are not well established, with very few studies that have performed a head-to-head comparison of GS and TGPs.^6,13–15^ Barriers to implementing clinical GS include challenges in interpreting results from GS, which have the potential to include less well-established disease genes and larger numbers of incidental findings.^12^

Understanding the molecular diagnostic yield of GS compared to TGPs in diverse patient populations is needed to support widespread clinical implementation of GS. The NYCKidSeq study aims to assess the understanding of genomic test results using a novel digital platform^16^ and to evaluate the diagnostic yield of GS and TGPs in diverse patient populations.^17^ The project is one of six studies funded as part of the Clinical Sequencing Evidence-Generating Research (CSER) 2 consortium.^18^ The present study focuses on outcomes related to the molecular diagnostic yield and concordance of GS and TGP testing in children and young adults with suspected genetic conditions from the NYCKidSeq study.

## MATERIALS AND METHODS

### Setting and study population

We recruited 650 probands and their families into the NYCKidSeq study, a randomized controlled trial (NCT03738098),^17^ between January 2019 and November 2020. Probands were recruited from the Mount Sinai Health System (N = 401) and the Montefiore Medical Center (N = 244) in New York City. Probands were aged ≤ 21 years at enrollment, with at least one English- or Spanish-speaking parent or legal guardian available to participate for completion of study surveys. Probands were patients receiving medical care at the participating health systems and were referred into the study by their healthcare providers. Individuals were eligible if they had neurologic (epilepsy/seizure disorder [epilepsy] or intellectual developmental disability/global developmental delay [IDD]), cardiac (congenital heart disease, cardiomyopathy, or cardiac arrhythmia), or immunologic (features of primary immunodeficiency) conditions, with a suspected underlying genetic cause. Individuals with previous genetic testing were eligible as long as their previous results were considered uninformative for their primary phenotype. Individuals with a known or likely (based on clinical features) molecular genetic diagnosis or with a previous bone marrow transplant were not eligible. Further details on recruitment, enrollment, and inclusion/exclusion criteria are described in Odgis *et al*.^17^ Parents/legal guardians completed a baseline survey at enrollment, in which they were asked to select the racial and/or ethnic category or categories that best described their child. Responses were then mapped into population groups (**eMethods 1**). This study was approved by the Icahn School of Medicine at Mount Sinai and the Albert Einstein College of Medicine Institutional Review Boards. All probands or parents/legal guardians provided written informed consent.

### Genetic testing

Blood and/or saliva samples were collected from the proband and their biological parent(s), if available. Genomic DNA was extracted from blood or saliva specimens using standard methods. Of the 650 probands enrolled, five withdrew from the study prior to genetic testing. TGPs were ordered for 644 probands at Sema4,^19^ and GS was ordered for 643 probands at the New York Genome Center (NYGC).^20^ All genetic testing was New York State approved and Clinical Laboratory Improvement Amendments (CLIA)-certified. TGPs consisted of a neurodevelopmental panel (447 genes), comprehensive immunodeficiency panel (250 genes), and/or comprehensive cardiovascular panel (240 genes) (**eTable 1**). For GS, either KAPA Hyper Prep kit (KAPA Biosystems) was used for sequencing on the Illumina HiSeq X instrument (samples submitted January 2019 to February 2020), or TruSeq DNA Nano or TruSeq DNA PCR-free library prep kits (Illumina, Inc.) were used for sequencing on the Illumina NovaSeq 6000 (samples submitted February 2020 onwards). Details about TGP testing and GS are in **eMethods 2** and **eMethods 3**. Sequence variants were classified according to standards from the American College of Medical Genetics and Genomics (ACMG).^21–23^ Variant interpretation and reporting for GS and TPG testing was performed independently by the two separate laboratories.

**Table 1.** Characteristics of 645 probands who underwent genetic testing in the NYCKidSeq study

| Characteristic | Probands, No. (%) |
| --- | --- |
| **Age, median (range)** | 9 years (2 months to 21 years) |
| **Female** | 247 (38.3) |
| **Male** | 398 (61.7) |
| **Population category** |  |
| American Indian, Native American, or Alaska Native | 1 (0.2) |
| Asian | 35 (5.4) |
| Black or African American | 105 (16.3) |
| Hispanic/Latino(a) | 328 (50.9) |
| Middle Eastern or North African/Mediterranean | 5 (0.8) |
| White or European American | 126 (19.5) |
| More than one population selected | 27 (4.2) |
| Other | 4 (0.6) |
| Prefer not to answer | 8 (1.2) |
| Unknown/none of these fully describe my child | 6 (0.9) |
| **Recruitment site** |  |
| Mount Sinai Health System | 401 (62.2) |
| Montefiore Medical Center | 244 (37.8) |
| **Primary phenotype** |  |
| Cardiac | 33 (5.1) |
| Immunologic | 38 (5.9) |
| Neurologic | 574 (89.0) |
| **Previous genetic testing** |  |
| Yes | 192 (29.8) |
| No | 453 (70.2) |

### Result interpretation

Case-level interpretation of genomic test result(s) was generated by a study genetic counselor (GC). The GC reviewed each variant, its respective laboratory classification, parental inheritance (if known), and the classic phenotype(s) associated with the gene(s) involved. A clinical interpretation was then assigned for each case as positive, likely positive, uncertain, or negative. A clinical interpretation of “positive” was achieved if all the following criteria were met: 1) variants classified as pathogenic or likely pathogenic (P/LP), 2) variants in genes associated with a condition consistent with the proband’s primary phenotype and/or family history, and 3) variants in allele states consistent with the inheritance pattern of the associated condition. A “likely positive” interpretation was achieved for P/LP variants in genes associated with a condition partially consistent with the proband’s primary phenotype (n = 8), variants of uncertain significance (VUS) in genes associated with a condition consistent with the primary phenotype (n = 6), mosaic results (n = 5), results with discordant variant interpretations including at least one P/LP interpretation (n = 4), and other cases (n = 3). Diagnosed cases were those with a positive or likely positive clinical interpretation resulting from either test modality. Discrepancies between the two testing modalities were noted, and GCs could seek input from an interpretation committee composed of study physicians with expertise in medical genetics (N.S.A.-H., G.A.D., B.D.G., J.M.G., and M.P.W.). Probands and biological parents could also opt into receiving secondary findings from GS, which included P/LP variants in 59 genes recommended for result return in the ACMG secondary findings (SF) v2.0 list.^24^

### Statistical analyses

Statistical analyses were performed from September 2021 to May 2022. The main outcome was the molecular diagnostic yield of GS and TGPs, defined as the proportion of probands for whom genetic testing yielded a positive or likely positive clinical interpretation. We used median (range) to describe continuous variables that were not normally distributed and proportions to describe categorical variables. Pearson’s chi-squared test was used to compare categorical variables. We used a fully paired study design to compare the diagnostic yield of TGP testing and GS in probands who underwent testing by both modalities, thereby fully controlling for sample-level covariates. McNemar’s test was used for all within sample dichotomous comparisons. Power analysis was performed to determine a minimum sample size of N = 45 was required for 80% power to surpass *P* < .05 using McNemar’s test; therefore, sub-groups with < 45 observations were not analyzed. All *P* values were from 2-tailed tests, and *P* < .05 was considered statistically significant. Statistical analyses were conducted using JMP Version 16 (SAS Institute Inc.).

## RESULTS

### Study population

We enrolled 650 probands and their families into the NYCKidSeq study between January 2019 and November 2020, and five withdrew prior to genetic testing. Among the 645 probands who underwent genetic testing, the median age at enrollment was 9 years (range 2 months to 21 years), and 38.3% were female (**Table 1**). The majority were Hispanic/Latino(a) (50.9%), White/European American (19.5%), and Black/African American (16.3%), based on surveys administered on enrollment. Most (89.0%) had a primary neurologic phenotype, and thirty-seven (5.7%) had more than one phenotype. There were 192 (29.8%) probands who had undergone genetic testing prior to enrollment. Among them, 31 (16.1%) had previous testing that included exome sequencing and another 104 (54.2%) had targeted gene panel testing (**eTable 2**). Neurodevelopmental panels were the most frequently ordered TGP in this study, and 29 (4.5%) probands had two panels ordered (**eTable 2**). In total, 642 probands had both TGP testing and GS; two had TGP testing only, and one had GS only (due to insufficient sample for GS or TGPs, respectively). The majority of cases had duo (34.2%) or trio (61.7%) analysis by GS (**eTable 2**).

**Table 2.** Overall diagnostic yield in 645 probands with suspected genetic conditions who underwent genetic testing in the NYCKidSeq study

| Variable | No./total No. | % (95% CI) | P value |
| --- | --- | --- | --- |
| **Age group** |  |  | .02 |
| < 3 years (infants/toddlers) | 21/73 | 28.8 (19.7 - 40.0) |  |
| 3 to 12 years (preschool/school age children) | 57/334 | 17.1 (13.4 - 21.5) |  |
| > 12 years (adolescents/young adults) | 35/238 | 14.7 (10.8 - 19.8) |  |
| **Population group** |  |  | .31<sup>a</sup> |
| American Indian, Native American, or Alaska Native | 0/1 | - |  |
| Asian | 3/35 | 8.6 (3.0 - 22.4) |  |
| Black or African American | 13/105 | 12.4 (7.4 - 20.0) |  |
| Hispanic/Latino(a) | 62/328 | 18.9 (15.0 - 23.5) |  |
| Middle Eastern or North African/Mediterranean | 1/5 | 20.0 (3.6 - 62.4) |  |
| White or European American | 25/126 | 19.8 (13.8 - 27.7) |  |
| More than one population selected | 5/27 | 18.5 (8.2 - 36.7) |  |
| Other | 1/4 | 25.0 (4.6 - 69.9) |  |
| Prefer not to answer | 2/8 | 25.0 (7.1 - 59.1) |  |
| Unknown/none of these fully describe my child | 1/6 | 16.7 (3.0 - 56.4) |  |
| **Recruitment site** |  |  | .001 |
| Mount Sinai Health System | 55/401 | 13.7 (10.7 - 17.4) |  |
| Montefiore Medical Center | 58/244 | 23.8 (18.9 - 29.5) |  |
| **Previous genetic testing** |  |  | .001 |
| Yes | 48/192 | 25.0 (19.4 - 31.6) |  |
| No | 65/453 | 14.3 (11.4 - 17.9) |  |
| **Primary phenotype** |  |  | .31 |
| Cardiac | 9/33 | 27.3 (15.1 - 44.2) |  |
| Immunologic | 7/38 | 18.4 (9.2 - 33.4) |  |
| Neurologic | 97/574 | 16.9 (14.1 - 20.2) |  |
| **Neurologic phenotype category** |  |  | <.001 |
| Epilepsy | 15/174 | 8.6 (5.3 - 13.7) |  |
| Epilepsy and intellectual developmental disability | 37/205 | 18.0 (13.4 - 23.9) |  |
| Intellectual developmental disability | 45/195 | 23.1 (17.7 - 29.5) |  |

<sup>a</sup> Chi squared test performed for population groups with ≥ 10 probands: Asian, Black or African American, Hispanic/Latino(a), White or European American, and More than one population

### Overall diagnostic yield of genetic testing

We reviewed clinical result interpretations in all 645 probands who underwent genetic testing (**eTable 3**). In total, 113 (17.5%) received a molecular diagnosis that fully or partially explained their phenotype, including 87 (13.5%) with positive and 26 (4.0%) with likely positive clinical interpretations. The remaining clinical interpretations were 373 (57.8%) uncertain and 159 (24.7%) negative. Among 643 probands who underwent GS, 503 opted to receive secondary findings, and 13 of these (2.6%) were found to have a P/LP variant in one of the 59 genes from the ACMG SF v2.0 list (**eTable 4**).^7^

**Table 3.**
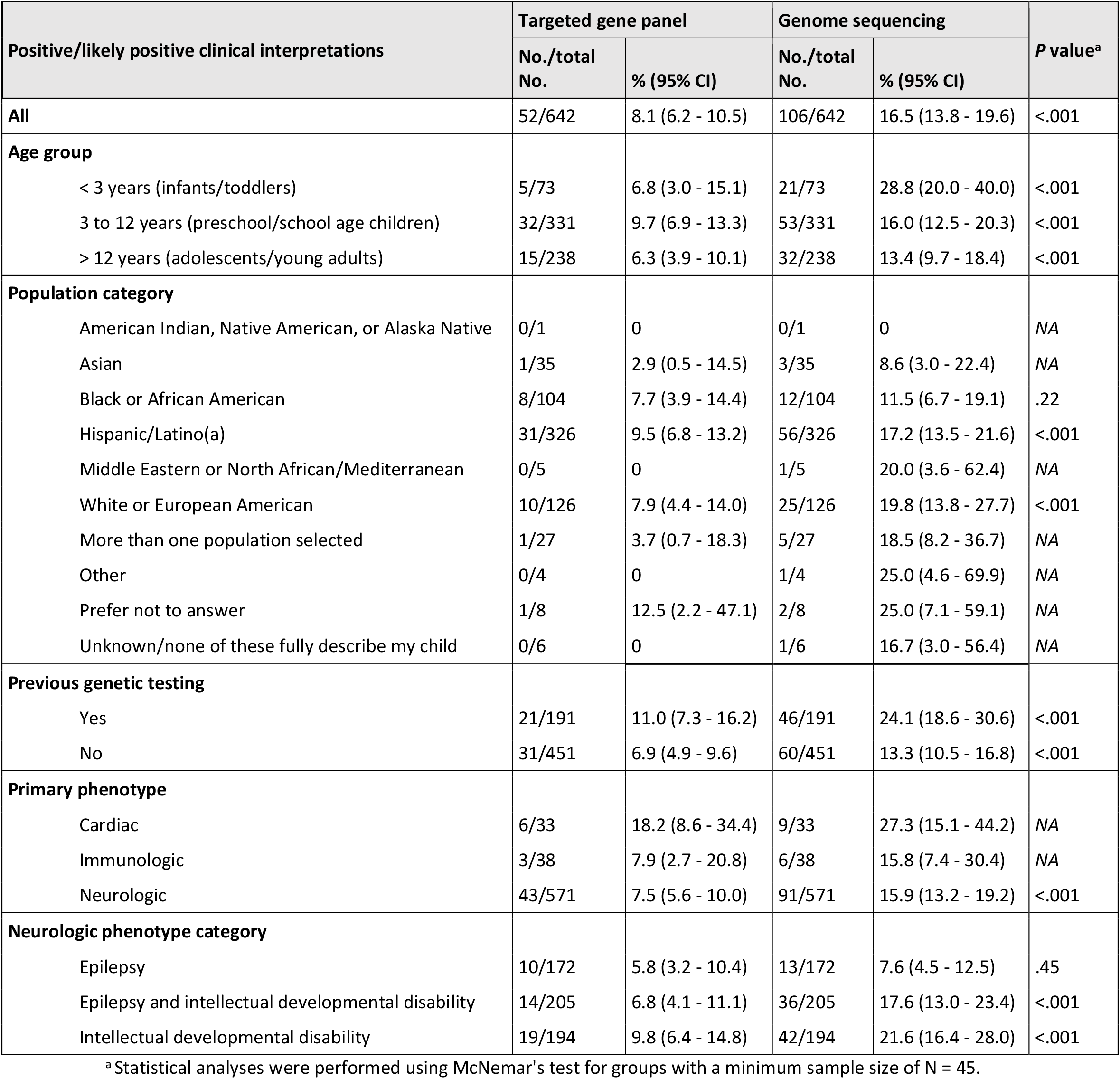
Molecular diagnostic yield by genetic test modality in 642 probands who underwent both targeted gene panel testing and genome sequencing in the NYCKidSeq study

Details of all 113 probands with a molecular diagnosis are in **eTable 5**. Their median age was 8 years (range 5 months to 21 years). Ten (8.8%) were found to have two variants associated with an autosomal recessive condition, including six homozygotes and four compound heterozygotes. Three (2.7%) had variants in two distinct genes that were each related to the proband’s phenotype; all three were considered to have multiple molecular diagnoses and a blended phenotype. Nine genes were implicated in two or more probands with neurologic conditions: *ARID1B* (N = 2), *CUX1* (N = 2), *KCNQ3* (N = 2), *KMT2A* (N = 3), *NAA15* (N = 2), *SCN1A* (N = 2), *SCN8A* (N = 6), *SETD5* (N = 2), and *SOX5* (N = 2). The variants identified in these genes were unique in each case.

The proportion of probands with positive or likely positive clinical interpretations is referred to from here on as the diagnostic yield. We assessed whether the diagnostic yield varied by age, population group, academic medical center, primary phenotype, or previous genetic testing (**Table 2**). Infants and toddlers (under 3 years) were more likely to receive a diagnosis than older children (age 3 to 12 years) or adolescents/young adults (28.8% *vs*. 17.1% and 14.7%, respectively; *P* = .02). Diagnostic yield was higher in probands who had genetic testing prior to enrollment compared to those who did not (24.9% *vs*. 14.4%; *P* = .001). Diagnostic yield was higher in probands recruited from the Montefiore Medical Center compared to those recruited from the Mount Sinai Health System (23.8% *vs*. 13.7%, *P* = .001). However, a larger proportion of probands from Montefiore had previous genetic testing (49.2% *vs*. 18.2%), and there was no difference in yield between the two sites when restricting to probands without previous genetic testing (18.6% for Montefiore *vs*. 12.8% for Mount Sinai, *P* = .12). Diagnostic yield did not vary across primary phenotypes (range 16.9% to 27.3%; *P* = .31). In probands with a primary neurologic phenotype, diagnostic yield was higher in probands with IDD or both epilepsy and IDD, compared to those with epilepsy alone (23.1% and 18.0% *vs*. 8.6%, respectively; *P* < .001).

### Diagnostic yield of genome sequencing versus targeted gene panel testing

Next, we evaluated whether the diagnostic yield varied by test modality among 642 probands who underwent both TGP testing and GS (**Table 3**). TGPs yielded 52 (8.1%) and GS yielded 106 (16.5%) molecular diagnoses (*P* < .001). We performed sub-analyses within population groups and primary phenotype category in groups with a minimum sample size of N = 45, which was required for 80% power to surpass *P* < .05 using McNemar’s test. Diagnostic yield was greater for GS compared to TGPs in self-reported Hispanic/Latino(a) (17.2% *vs*. 9.5%, *P* < .001) and White/European American (19.8% *vs*. 7.9%, and *P* < .001) population groups, but not in the self-reported Black/African American group (11.5% *vs*. 7.7%, *P* = .22) (**Figure 1A**). Diagnostic yield was greater for GS compared to TGPs in probands with or without previous genetic testing (*P* < .001 for both). In probands with a primary neurologic phenotype, diagnostic yield was greater for GS compared to TGP in those with IDD or both epilepsy and IDD (*P* < .001 for both), but not in probands with epilepsy alone (*P* = .45).

**Figure 1.**
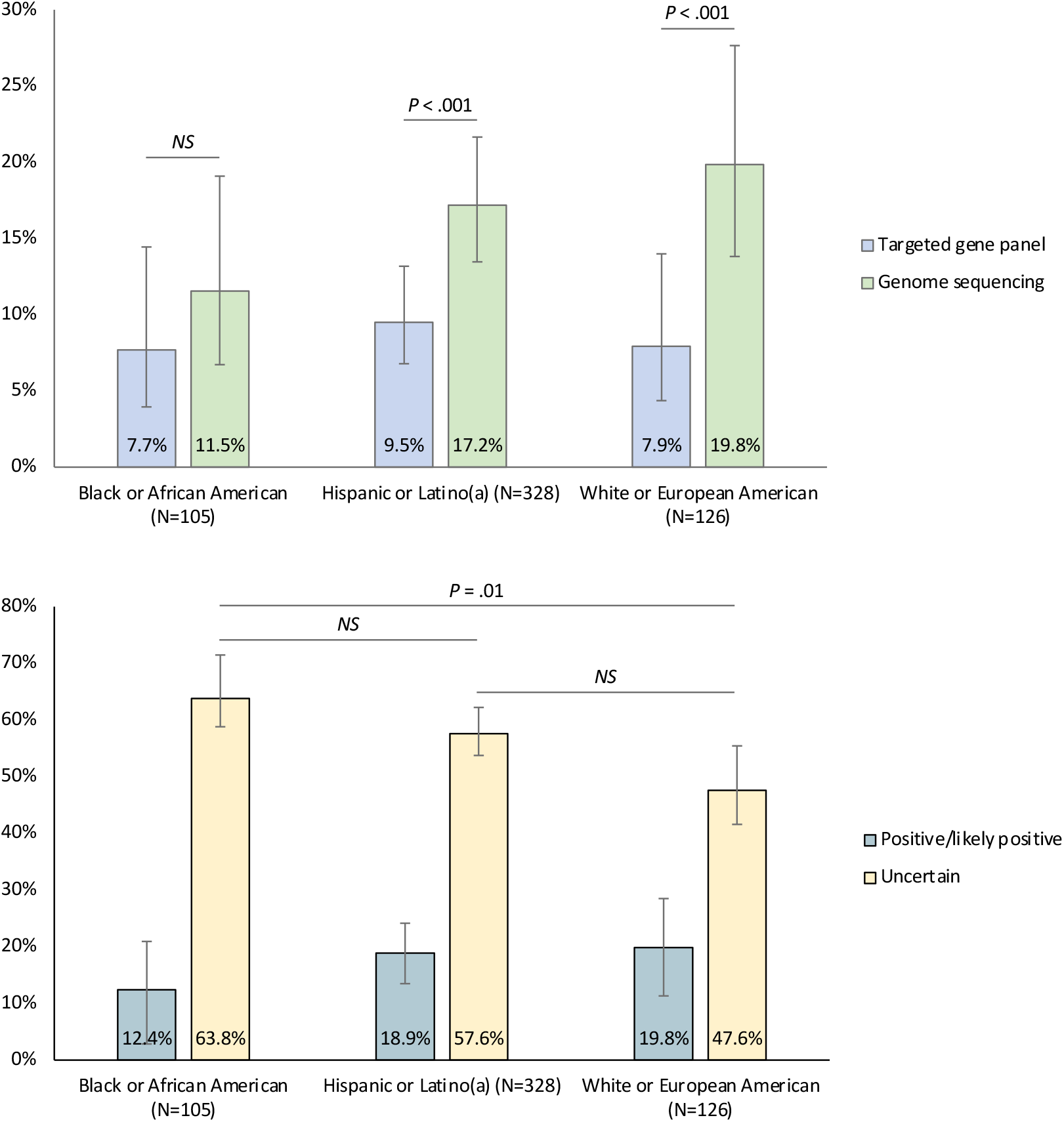
**A**. Molecular diagnostic yield (proportion of positive/likely positive clinical interpretations) of targeted gene panel (TGP) testing and genome sequencing (GS) across the largest population groups by self-report. Diagnostic yield was higher for GS compared to TGP testing in self-reported Hispanic/Latino(a) and White/European American but not in Black/African American population groups. **B**. Rates of positive/likely positive and uncertain clinical interpretations across population groups. Rates of positive/likely positive clinical interpretations (i.e. diagnostic yield) did not vary significantly across groups. Rates of uncertain clinical interpretations were higher in self-reported Black/African American than White/European American population groups. Error bars represent 95% confidence intervals. *NS*, not significant.

We hypothesized that differences in diagnostic yield between GS and TGPs across population groups could be due to differences in the number of inconclusive results obtained in each group. Therefore, we compared the proportion of probands receiving an uncertain clinical interpretation across population groups (**eTable 6**). We found a higher proportion of uncertain clinical interpretations in Black/African American (63.8%) compared to White/European American (47.6%; *P* = .01) probands (**Figure 1B**).

### Concordance of genome sequencing and targeted gene panel testing

Among the 113 probands who received a molecular diagnosis, there were 45 diagnoses from both TGP and GS, 7 TGP-only diagnoses, and 61 GS-only diagnoses (**Figure 2** and **eTable 5**). The majority of variants reported by both TGP and GS were SNVs (n = 20), small indels (n = 15), and intronic variants (n = 9). Nineteen probands had at least one CNV (*i*.*e*., an unbalanced genomic gain or loss greater than 50 base pairs), 17 of which were reported by GS alone. In four of the 17 cases with GS-only reported CNVs, TGPs partially identified the CNV and recommended follow-up by high-resolution array (**eTable 7**). Eight probands were found to have a mosaic variant, six of which were reported by GS alone. Two of the mosaic variants (NM_000548.5 (*TSC2*) c.1257+5G>A and NM_181552.4 (*CUX1*) c.2014C>T) were reported by GS only after expanding from singleton to duo or trio re-analysis (see **eMethods 3**).

**Figure 2.**
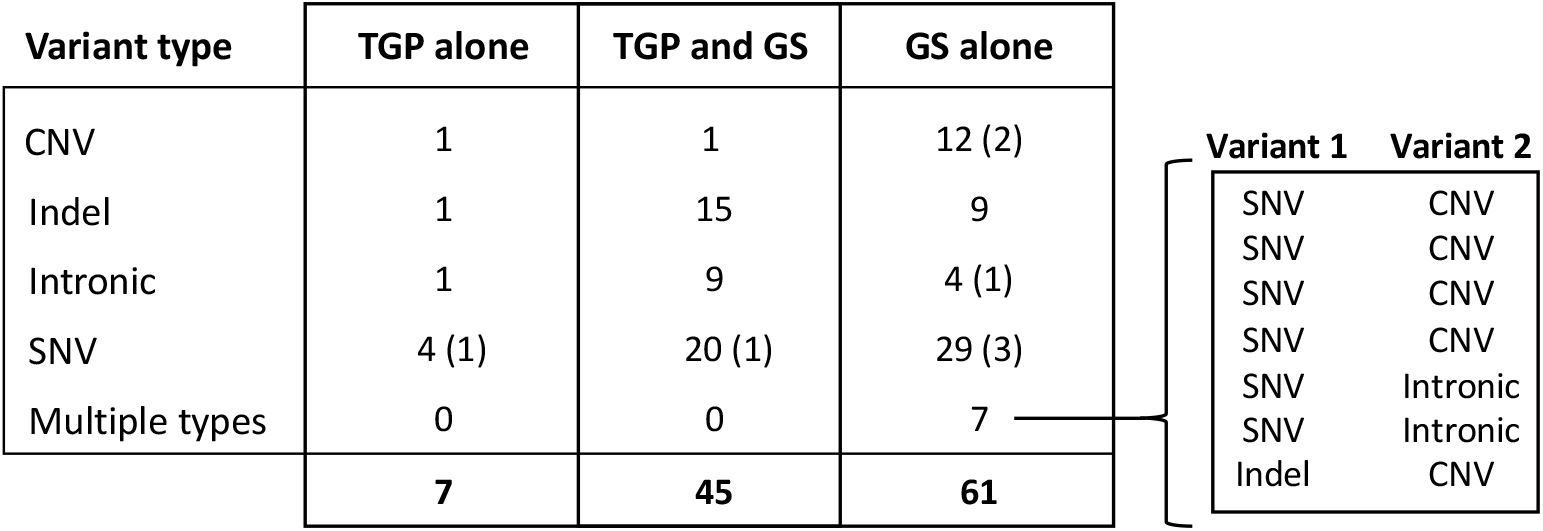
Concordance between targeted gene panel (TGP) testing and genome sequencing (GS). The number and types of variants identified by TGP testing, GS, or both test modalities are shown. Eight mosaic variants were identified, including six by GS alone, one by TGP testing alone, and one by both modalities; these are indicated in parentheses for each test modality and variant type. There were seven probands for whom two distinct variants were identified (*i*.*e*., multiple types). CNV, copy number variant; Indel: insertion/deletion; SNV; single nucleotide variants.

Of the 61 GS-only diagnoses, 38 were in genes not present on the TGPs used in the NYCKidSeq study. The 23 GS-only diagnoses that were in genes present on the panels were considered discrepant results between the two modalities (**eTable 7**). Of these, 6 were variants reported by TGP but classified as VUS. One variant (NM_001080517.3 (*SETD5*) c.972T>G) was later updated to P/LP as a result of communication from the study team with the TGP testing laboratory. Seventeen GS-only diagnoses were variants not reported by TGPs, despite the gene being present on the panel(s). These included eight probands with CNVs, four with intronic variants, two with indels, two with SNVs, and one with both a SNV and intronic variant. One indel, NM_001372044.1 (*SHANK3*) c.4634dup, was not detected by TGP because the laboratory was reporting only CNVs in *SHANK2* and *SHANK3* at the time of analysis; TGP testing was later modified to include sequencing of these genes (**eTable 1**). The seven TGP-only diagnoses included four SNVs (one of which was mosaic), one CNV, one indel, and one intronic variant. Five of the seven TGP-only diagnoses were variants reported by GS but classified as VUS. The remaining two variants, NM_005249.4 (*FOXG1*) c.216del and NM_001008537.2 (*NEXMIF*) c.2030C>A (mosaic), were not detected by the GS bioinformatic pipeline due to lack of adequate sequence coverage at variant sites. All TGP-only diagnoses were considered discrepant results between the two modalities. Therefore, a total of 30 diagnosed cases (26.5%) had discrepancies, which mainly included variants detected by one modality but not the other or differences in variant classifications (**eTable 7**). Given that differences in variant detection is a separate issue from differences in variant classification, we evaluated the concordance of TGP testing and GS on variant detection only. For this, we considered the 11 cases with variant classification differences as being detected by both test modalities, resulting in 56 diagnoses from both TGP and GS, 2 TGP-only diagnoses, and 55 GS-only diagnoses (**eFigure 1**).

Differences in diagnostic yield between GS and TGPs may be partially explained by GS analysis of duos or trios compared to singleton analysis by TGP testing, or by the selection of gene panels for TGP testing, which can vary by gene content. To evaluate the first possibility, we compared the diagnostic yield of TGPs and GS in 343 cases for which GS was initially performed for probands only (**eMethods 3** and **eTable 3**). In this subset, diagnostic yield for GS remained higher than that for TGP testing (15.5% *vs*. 6.1%, respectively; P < .001). Next, we performed an *in silico* analysis using seven commercially available gene panels from Ambry^25^ and Invitae^26^ commonly ordered for the evaluation of neurologic, immunologic, and cardiac phenotypes (**eMethods 4** and **eTable 8**). Seventy of the 113 molecular diagnoses (61.9%) were in a gene present on at least one panel, including 25 of the 61 GS-only diagnoses (41.0%; **eTable 9**). Using GS in combination with all seven commercially available panels, we could maximally achieve 111 of the 113 molecular diagnoses from NYCKidSeq, including 65 diagnoses from both TGPs and GS, 5 TGP-only diagnoses, and 41 GS-only diagnoses. The maximum diagnostic yield for this combination of TGPs was 10.9%.

## DISCUSSION

This study from the NYCKidSeq clinical trial investigated the utility of GS as a first-line diagnostic test. It included a large, racially and ethnically diverse pediatric and young adult patient population with heterogeneous clinical presentations, enrolled from across medical specialties in two academic medical centers. We used a fully paired study design to compare GS and TGP test results, and the main outcome of interest was the proportion of probands who received a molecular diagnosis by each test modality. Our findings demonstrate an overall molecular diagnostic yield of 17.5% in 645 children and young adults with suspected genetic conditions. Comparing the two test modalities, we found that GS yielded a diagnosis in 16.5% of probands, while TGP testing using panels of 240 to 447 genes yielded a diagnosis in 8.1%.

The overall diagnostic yield in the NYCKidSeq study is in line with other studies, which report diagnostic yields from clinical GS ranging from 14% - 41%, depending on the study population, selection criteria, and disease area.^2,8,27–30^ Diagnostic yield in our study was highest in children < 3 years of age (28.8%) compared to older age groups and was higher in probands who had had previous non-informative clinical genetic testing (24.9%) compared to those who had not (14.4%). This may reflect that probands with previous genetic testing were those with features most suggestive of an underlying genetic etiology. We found multiple molecular diagnoses in 2.7% of cases in which genetic testing was informative, which is consistent with retrospective studies reporting 4.6% to 4.9% of diagnosed cases having multiple molecular diagnoses and a blended phenotype.^31,32^ Among 574 probands with a primary neurologic phenotype, the diagnostic yield was 16.9% and was higher in probands with IDD (23.1%) or both epilepsy and IDD (18.0%). Previous studies with smaller numbers of children have reported the molecular diagnostic yield of genetic testing in pediatric neurodevelopmental diseases to be around 30% - 40%, which is higher than what we observed, and may reflect differences in inclusion criteria, which were relatively broad in NYCKidSeq.^33–35^

We compared the diagnostic yield and concordance of GS and TGP in 642 probands who had both types of tests and found that less than 40% (45 of 113) of diagnoses were achieved by both test modalities. GS-only diagnoses included structural variants and non-exonic sequence variants; however, the majority of GS-only diagnoses were exonic SNVs, most of which were in genes not present on the TGPs. GS also identified several variants (including CNVs and intronic variants) in genes present on the panels used in the study. Seventeen of the 19 CNVs detected in probands with neurologic, immunologic and/or cardiac phenotypes were reported by GS alone. In four of these cases, TGP partially identified the CNV in question and recommended additional testing using high-resolution chromosomal microarray as a next step to delineate the full size of the CNV. The majority of mosaic variants (six of eight) were identified by GS alone, including five that were in genes present on the panels used in the study. Details of mosaic variants detected in the NYCKidSeq study are discussed separately.^36^ Among probands with primary neurologic phenotypes, only those with epilepsy and no IDD had similar diagnostic yields between TGP testing (5.8%) and GS (7.6%), suggesting that TGPs could be recommended as a first-line test in isolated epilepsy cases. The gene content of specific panels used in our study did not fully account for diagnostic yield differences between GS and TGP testing. An *in silico* analysis replacing the TGPs used in NYCKidSeq with a combination of seven commercially available panels showed a maximum yield of 10.9%. Therefore, we can estimate the diagnostic yield of clinical TGP testing to be approximately 8 to 11%, depending on choice of panel(s). In this study, GS performed in singletons still had a higher yield than TGP testing. If confirmed in other studies, perhaps those examining other clinical contexts, this might have a significant impact on resource utilization for testing with GS.

While the overall diagnostic yield did not differ across population groups in the NYCKidSeq study, diagnostic yield of GS was significantly greater than that of TGP testing in self-reported Hispanic/Latino(a) (17.2% *vs*. 9.5%) and White/European American (19.8% *vs*. 7.9%) population groups, but not in the self-reported Black/African American (11.5% *vs*. 7.7%) group. We hypothesized that this could be due to increased rates of inconclusive results in individuals self-identifying as Black/African American. Indeed, we found a higher proportion of uncertain clinical interpretations in Black/African American probands (63.8%) compared to those who were Hispanic/Latino(a) or White/European American (57.6% and 47.6%, respectively). Recent studies have similarly reported lower diagnostic yields of genetic testing and higher rates of inconclusive results in Black children and adults with cardiomyopathy, and in Black children with sensorineural hearing loss.^37,38^ It is plausible that GS in our study did not improve the diagnostic yield in Black/African American individuals because of greater uncertainty in interpreting genomic variants in this population group. Such inequities in clinical genetic test interpretation are related to lower rates of genetic and genomic research studies that include underrepresented populations, an issue that has garnered more attention in recent years, but that remains an ongoing problem.^39^

There are limitations to this study. As the majority of probands had a primary neurologic phenotype, diagnostic yields reported in this study may not be generalizable to other types of pediatric conditions. Despite the racial and ethnic diversity of the cohort, there was low representation of certain population groups, and only the three largest groups were sufficiently powered to evaluate differences in diagnostic yield between GS and TGP within those groups. GS and TGP testing were performed at two separate commercial labs, so inter-lab variability in variant analysis and result interpretation could not be accounted for in evaluating concordance between the two testing modalities. For example, eleven of the discrepant results between GS and TGP involved variant classification differences, including five P/LP variants from TGP reported as VUS by GS, and six P/LP variants from GS reported as a VUS by TGP. Recently reported rates of VUS results from gene panels and exome/genome sequencing range from 6.0% for panel tests of 2-10 genes to 76.2% for panel tests >200 genes, and 22.5% for exome/genome sequencing.^40^ Our relatively high rates of uncertain clinical interpretations in NYCKidseq (57.8%) may be due to the large panel sizes used (all >200 genes) as well as demographic differences in our study population. Our study did not address turnaround time, which is an important consideration for implementation of GS as a first-line diagnostic test. GS and TGP turnaround times were not informative in the present study due to sample processing workflows that were not reflective of typical clinical and laboratory practice.

Establishing a definitive diagnosis for individuals with conditions of presumed genetic origin can guide medical care, improve understanding of disease progression, and inform recurrence risk for reproductive planning. The evaluation of pediatric genetic conditions today often involves multiple clinical, imaging, and laboratory tests, which can prolong the diagnostic odyssey and incur substantial costs. Our study demonstrates the utility of GS as a diagnostic test in children and young adults with suspected genetic conditions, and provides strong evidence to support its use early in the diagnostic trajectory. However, we note that the lack of improvement in diagnostic yield with GS coupled with increased rates of uncertainty in Black/African American individuals indicate that there is much more work to do to ensure that GS implementation in clinical care benefits all populations and does not widen health disparities.

## Data Availability

De-identified data for this study will be shared in secure, access-restricted scientific research databases called NHGRI Analysis Visualization and Informatics Lab-space (AnVIL) and the Database of Genes and Phenotypes (dbGaP) at the National Institutes of Health (dbGaP accession number phs002337.v1.p1). Interpretations of the clinical significance of variants from genetic testing have been submitted to the ClinVar database at the National Institutes of Health under the study name NYCKidSeq.

## Data Availability

De-identified data for this study will be shared in secure, access-restricted scientific research databases called NHGRI Analysis Visualization and Informatics Lab-space (AnVIL) and the Database of Genes and Phenotypes (dbGaP) at the National Institutes of Health (dbGaP accession number phs002337.v1.p1). Interpretations of the clinical significance of variants from genetic testing have been submitted to the ClinVar database at the National Institutes of Health under the study name ‘NYCKidSeq’.

## Acknowledgments

The NYCKidSeq Program (U01HG0096108; Principal Investigators Kenny, Wasserstein, Gelb and Horowitz) is supported by the Clinical Sequencing Evidence-Generating Research (CSER) consortium, which is funded by the National Institutes of Health (NIH) National Human Genome Research Institute, National Institute for Minority Heath and Health Disparities of the National Institutes of Health and the National Cancer Institute. The contents of this paper are solely the responsibility of the authors and do not necessarily represent the official views of the NIH. We thank the providers who referred patients to the study for facilitating recruitment, and the Mount Sinai Genomics Stakeholder Board for their thoughtful involvement in all aspects of the study. We thank and are grateful to the patients and their families who contributed to this study.

## Author Contributions

Dr. Abul-Husn had full access to all of the data in the study and takes responsibility for the integrity of the data and the accuracy of the data analysis. Dr. Kenny and Dr. Gelb oversaw all aspects of the study and are co-senior authors.

Conceptualization: Greally, Horowitz, Wasserstein, Kenny, Gelb; Methodology: Abul-Husn, Kenny, Gelb; Data curation and investigation: Abul-Husn, Marathe, Kelly, Bonini, Sebastin, Odgis, Abhyankar, Brown, Di Biase, Gallagher, Guha, Ioele, Okur, Ramose, Rodriguez, Rehman, Thomas-Wilson, Edelmann, Zinberg, Diaz, Greally, Jobanputra, Suckiel, Horowitz, Wasserstein, Kenny, Gelb; Formal analysis: Abul-Husn, Kenny; Funding acquisition: Kenny, Gelb, Horowitz, Wasserstein; Project administration: Kelly, Ramos, Rodriguez; Supervision: Kenny, Gelb, Horowitz, Wasserstein; Writing – original draft: Abul-Husn, Kenny, Gelb; Writing – review and editing: Abul-Husn, Marathe, Kelly, Bonini, Greally, Jobanputra, Suckiel, Wasserstein, Kenny, Gelb.

## Ethics Declarations

Disclosures: Dr. Abul-Husn is an employee and equity holder of 23andMe; serves as a scientific advisory board member for Allelica; received personal fees from Genentech, Allelica, and 23andMe; received research funding from Akcea; and was previously employed by Regeneron Pharmaceuticals. Dr. Kenny received personal fees from Illumina, 23andMe, Allelica, and Regeneron Pharmaceuticals, received research funding from Allelica, and serves as a scientific advisory board member for Encompass Bio, Foresite Labs, and Galateo Bio. All other authors declare they have no disclosures to report.

Ethics Approval and Consent: This study was approved by the Icahn School of Medicine at Mount Sinai and the Albert Einstein College of Medicine Institutional Review Boards. All probands or parents/legal guardians provided written informed consent.

