## Supplemental data for "Molecular diagnostic yield of genome sequencing versus targeted gene panel testing in racially and ethnically diverse pediatric patients"

**Supplementary Online Content**

**eMethods 1.** Mapping population groups

**eMethods 2.** Targeted gene panel testing

**eMethods 3.** Genome sequencing

**eMethods 4.** *In silico* analysis of diagnostic yield

**eReferences**

**eTable 2.** Proband phenotypes and targeted gene panels ordered in the NYCKidSeq study

**eTable 3.** Clinical interpretation of genetic testing in 645 probands at the time of initial and final genome sequencing reports

**eTable 4.** Secondary findings reported in 13 of the 503 NYCKidSeq participants who opted to receive secondary findings

**eTable 6.** Uncertain clinical interpretations across population groups in the NYCKidSeq study

**eTable 7.** Discrepancies observed between targeted gene panel and genome sequencing reports among probands with a molecular diagnosis

**eTable 9.** *In silico* analysis of diagnostic yield using a combination of seven commercially available targeted gene panels

**eFigure 1.** Concordance of detected variants between targeted gene panel (TGP) testing and genome sequencing (GS) in 113 probands who received a molecular diagnosis.

**eMethods 1.** Mapping population groups

Parents/legal guardians completed a baseline survey (77% in English and 23% in Spanish) at enrollment, in which they were asked to select the racial and/or ethnic category or categories that best described their child.[1] Categories were: American Indian, Native American, or Alaska Native; Asian; Black or African American; Native Hawaiian/Pacific Islander; White or European American; Middle Eastern or North African/Mediterranean; Hispanic/Latino(a); prefer not to answer; unknown/none of these fully describe my child; and other category. Individuals who selected multiple categories including "Hispanic/Latino(a)" were designated as "Hispanic/Latino(a)". All other individuals who selected multiple categories were designated as "more than one population selected".

**eMethods 2.** Targeted gene panel testing

Clinical targeted gene panel (TGP) testing was performed at Sema4.[2] Genomic DNA was extracted from the probands' blood or saliva specimens using standard methods. The three TGPs used in the study consisted of a neurodevelopmental panel (447 genes), a comprehensive immunodeficiency panel (250 genes), and a comprehensive cardiovascular panel (240 genes) from Sema4 (**eTable 1**). One or more of the panels were ordered based on the participant's phenotype. Agilent SureSelectTM QXT DNA library prep kits with a custom capture library were used, which included the exonic regions of genes on the panel, +/- 5 bps at the exon/intron junctions, and previously reported variants in non-coding regions within the targeted genes. Illumina NovaSeq platform (Xp workflow) with 100-bp paired-end reads was used to sequence proband samples. Reference build GRCh37/hg19 was used for alignment and variant calling. A minimum coverage of 20X was established and bioinformatic algorithms designed and validated by the laboratory were used to analyze the sequenced data. Genomic DNA was analyzed by next-generation sequencing, copy number variant analysis, and fragile X CGG repeat analysis, as appropriate. Confirmatory methods used included multiplex ligation-dependent probe amplification (MLPA), exon array, quantitative PCR, and Sanger sequencing. Variants were classified according to interpretation guidelines from the American College of Medical Genetics and Genomics (ACMG) and the Association for Medical Pathology (AMP).[3]

**eMethods 3.** Genome sequencing

Clinical genome sequencing was performed at the New York Genome Center (NYGC).[4] Genomic DNA was extracted from blood or saliva specimens using standard methods, or DNA extracted at Sema4 was shipped to NYGC. Proband-only analysis was performed for samples submitted from January 2019 to February 2020. KAPA Hyper Prep kit (KAPA Biosystems) was used for sequencing on Illumina HiSeq X instrument (Illumina, CA). DNA sequences from the proband samples were aligned to human genome build 37 (GRCh37/hg19) reference sequence. Reportable variants were confirmed and segregation analysis performed using an orthogonal method such as Sanger sequencing or SNP microarray.

During the course of the study, Illumina provided reagents allowing for additional sequencing of parental samples. Therefore, for samples submitted after February 2020, singleton (proband-only), duo, or trio analysis was completed based on availability of parental samples. TruSeq DNA Nano or TruSeq DNA PCR-free library preparation kits were used for sequencing on the Illumina NovaSeq 6000 instrument (Illumina, CA). DNA sequences from the proband and available parental samples were aligned to human genome build 38 (GRCh38/hg38) reference sequence. Duo or trio re-analysis was also performed for 146 unsolved cases that were previously completed as a proband-only analysis. For this, GS was performed on parental samples using Illumina NovaSeq 6000 instrument (Illumina, CA). DNA sequences from the proband and parental samples were aligned to human genome build 38 (GRCh38/hg38) reference sequence and reanalysis completed as a duo or trio sample. Variants aligned to GRCh37/hg19 were lifted to GRCh38/hg38 for consistency in this manuscript.

All genomes were sequenced with 150 base pair paired-end reads to at least 30X +/- 3X mean coverage, and a minimum of 85% of bases are sequenced to at least 20X coverage. Genomes were analyzed and interpreted using a phenotype-directed approach. Variants were classified using variant interpretation guidelines from the ACMG, AMP, and the Clinical Genome Resource (ClinGen).[3,5,6] Reported variants (except large CNVs) were validated using Variant Validator.

A secondary findings analysis was offered to each individual tested as part of the family-based analysis (*e.g.*, proband and biological parents as part of a trio test) in accordance with the ACMG recommendations for the return of secondary findings. For individuals who opted in to receive secondary findings analysis, a targeted search was conducted for pathogenic or likely pathogenic variants in the following genes:

*ACTA2*, *ACTC1*, *APC*, *APOB*, *ATP7B*, *BMPR1A*, *BRCA1*, *BRCA2*, *CACNA1S*, *COL3A1*, *DSC2*, *DSG2*, *DSP*, *FBN1*, *GLA*, *KCNH2*, *KCNQ1*, *LDLR*, *LMNA*, *MEN1*, *MLH1*, *MSH2*, *MSH6*, *MUTYH*, *MYBPC3*, *MYH11*, *MYH7*, *MYL2*, *MYL3*, *NF2*, *OTC*, *PCSK9*, *PKP2*, *PMS2*, *PRKAG2*, *PTEN*, *RB1*, *RET*, *RYR1*, *RYR2*, *SCN5A*, *SDHAF2*, *SDHB*, *SDHC*, *SDHD*, *SMAD3*, *SMAD4*, *STK11*, *TGFBR1*, *TGFBR2*, *TMEM43*, *TNNI3*, *TNNT2*, *TP53*, *TPM1*, *TSC1*, *TSC2*, *VHL*, *WT1*

**eMethods 4.** *In silico* analysis of diagnostic yield

We performed an *in silico* analysis to estimate the diagnostic yield from commercially available TGPs not used in the NYCKidSeq study. We selected a set of neurodevelopmental, immunodeficiency, and cardiovascular TGPs from AmbryGenetics[7] and Invitae[8] that are routinely used in clinical practice by medical geneticists to evaluate patients with neurologic, immunologic, and cardiac phenotypes in the two academic medical centers participating in NYCKidSeq. The TGPs were: Ambry NeurodevelopmentNext (202 genes), Ambry EpilepsyNext (124 genes), Invitae Primary Immunodeficiency panel (429 genes), Ambry RhythmNext (42 genes), Ambry HCMNext (30 genes), Ambry TAADNext (35 genes), and Invitae Congenital Heart Disease panel (55 genes). Information about the genes included in these TGPs was accessed online from the two laboratory websites in April 2022 (**eTable 2**).

**eTable 2.** Proband phenotypes, previous genetic testing, targeted gene panels ordered, and genome sequencing performed in the NYCKidSeq study

| **Phenotype(s), N = 645** | **Probands, No. (%)** |
|---|---|
| Cardiac | 27 (4.2) |
| Immunologic | 27 (4.2) |
| Neurologic | 554 (85.9) |
| Cardiac and immunologic | 2 (0.3) |
| Cardiac and neurologic | 15 (2.3) |
| Immunologic and neurologic | 18 (2.8) |
| Cardiac, immunologic, and neurologic | 2 (0.3) |
| **Previous genetic testing, N = 192** | **Probands, No. (%)** |
| Including exome sequencing | 31 (16.1) |
| Including targeted gene panel (no exome sequencing) | 104 (54.2) |
| Other (no exome sequencing or targeted gene panel) | 56 (29.2) |
| Unknown | 1 (0.5) |
| **Targeted gene panel(s) ordered, N = 644** | **Probands, No. (%)** |
| Cardiovascular | 28 (4.3) |
| Immunodeficiency | 29 (4.5) |
| Neurodevelopmental | 558 (86.6) |
| Cardiovascular and immunodeficiency | 3 (0.5) |
| Cardiovascular and neurodevelopmental | 12 (1.9) |
| Immunodeficiency and neurodevelopmental | 14 (2.2) |
| **Genome sequencing performed, N = 643** | **Probands, No. (%)** |
| Singleton | 26 (4.0) |
| Duo | 220 (34.2) |
| Trio | 397 (61.7) |

**eTable 3.** Clinical interpretation of genetic testing and diagnostic yield of targeted gene panel testing and genome sequencing at the time of initial (proband-only) and final genome sequencing reports

| Clinical Interpretation | Initial proband-only analysis (N = 343), No. (%) | Final analysis (N = 645), No. (%) |
| --- | --- | --- |
| Positive | 44 (12.8) | 87 (13.5) |
| Likely positive | 12 (3.5) | 26 (4.0) |
| Uncertain | 184 (53.6) | 373 (57.8) |
| Negative | 103 30.0) | 159 (24.7) |
| **Positive and likely positive clinical interpretations by test modality** | **Initial proband-only analysis (N = 343), No. (%)** | **Final analysis (N = 642), No. (%)** |
| Targeted gene panel | 21 (6.1) | 52 (8.1) |
| Genome sequencing | 53 (15.5) | 106 (16.5) |

**eTable 4.** Secondary findings reported in 13 of the 503 NYCKedSeq participants who opted to receive secondary findings

| Age Group | Sex | Population group | Transcript ID | Gene (HGNC ID) | Genomic change (HGVS.g; GRCh38) | Nucleotide change (HGVS.c) | Amino acid change (HGVS.p) | Variant classification | Parental Inheritance |
|---|---|---|---|---|---|---|---|---|---|
| 3 to 12 years | Male | White or European American | NM_000038.6 | *APC* (HGNC:583) | g.112839514T>A | c.3920T>A | p.(Ile1307Lys) | Pathogenic | Maternal |
| >12 years | Male | White or European American | NM_000038.6 | *APC* (HGNC:583) | g.112839514T>A | c.3920T>A | p.(Ile1307Lys) | Pathogenic | Maternal |
| 3 to 12 years | Male | Unknown/none of these fully describe my child | NM_000059.4 | *BRCA2* (HGNC:1101) | g.32337185A>T | c.2830A>T | p.(Lys944Ter) | Pathogenic | Unknown |
| 3 to 12 years | Female | Hispanic/Latino(a) | NM_000059.4 | *BRCA2* (HGNC:1101) | g.32339497T>G | c.5142T>G | p.(Tyr1714Ter) | Likely pathogenic | Unknown |
| 3 to 12 years | Male | Hispanic/Latino(a) | NM_000527.5 | *LDLR* (HGNC:6547) | g.11107436G>A | c.862G>A | p.(Glu288Lys) | Pathogenic | Maternal |
| 3 to 12 years | Male | Multiple selected | NM_000527.5 | *LDLR* (HGNC:6547) | g.11116874G>A | c.1721G>A | p.(Arg574His) | Likely pathogenic | Maternal |
| <3 years | Male | Hispanic/Latino(a) | NM_000257.4 | *MYH7* (HGNC:7577) | g.23431584C>T | c.732+1G>A | | Pathogenic | Paternal |
| 3 to 12 years | Male | Multiple selected | NM_020975.6 | *RET* (HGNC:9967) | g.43114598G>C | c.1998G>C | p.(Lys666Asn) | Pathogenic | Maternal |
| 3 to 12 years | Male | Hispanic/Latino(a) | NM_003000.3 | *SDHB* (HGNC:10681) | g.17023975G>A | c.640C>T | p.(Gln214Ter) | Likely pathogenic | Maternal |
| 3 to 12 years | Male | Multiple selected | NM_003001.5 | *SDHC* (HGNC:10682) | g.161314406A>G | c.1A>G | p.(Met1?) | Pathogenic | Unknown |
| <3 years | Female | Hispanic/Latino(a) | NM_000363.5 | *TNNI3* (HGNC:11947) | g.55154172C>T | c.407G>A | p.(Arg136Gln) | Likely pathogenic | Unknown |
| >12 years | Male | Asian | NM_001276345.2 | *TNNT2* (HGNC:11949) | g.201364357G>A | c.430C>T | p.(Arg144Trp) | Likely pathogenic | Maternal |
| 3 to 12 years | Male | Hispanic/Latino(a) | NM_001018005.2 | *TPM1* (HGNC:12010) | g.63062598C>T | c.725C>T | p.(Ala242Val) | Likely pathogenic | Unknown |

**eTable 6.** Uncertain clinical interpretations across population groups in the NYCKidSeq study

| Population category | No./total No. | % (95% CI) |
|---|---|---|
| American Indian, Native American, or Alaska Native | 0/1 | - |
| Asian | 25/35 | 71.4 (54.9 - 83.7) |
| Black or African American | 67/105 | 63.8 (54.3 - 72.4) |
| Hispanic/Latino(a) | 189/328 | 57.6 (52.2 - 62.9) |
| Middle Eastern or North African/Mediterranean | 3/5 | 60.0 (23.1 - 88.2) |
| White or European American | 60/126 | 47.6 (39.1 - 56.3) |
| More than one population selected | 17/27 | 63.0 (44.2 - 78.5) |
| Other | 3/4 | 75.0 (30.1 - 95.4) |
| Prefer not to answer | 4/8 | 50.0 (21.5 - 78.5) |
| Unknown/none of these fully describe my child | 5/6 | 83.3 (43.6 - 97.0) |

**eTable 7.** Discrepancies observed between targeted gene panel and genome sequencing reports among probands with a molecular diagnosis

| Gene or chromosomal coordinates | Variant type | Mosaic | Genetic test modality | Clinical interpretation | Type of discrepancy |
|---|---|---|---|---|---|
| *AIRE* (HGNC:360*), AIRE* (HGNC:360) | Intronic (+ SNV) | No | GS alone | Likely positive | GS identified a P/LP variant. TGP did not and gene of interest was analyzed. |
| *CARD14* (HGNC:16446) | SNV | No | TGP alone | Positive | Variant classifications differed between GS and TGP reports. |
| *FOXG1* (HGNC:3811) | Indel | No | TGP alone | Positive | TGP identified a P/LP variant. GS did not. |
| *GABRB2* (HGNC:4082) | SNV | Yes | GS alone | Likely positive | GS identified a P/LP variant. TGP did not and gene of interest was analyzed. |
| *GNAO1* (HGNC:4389) | Intronic | No | GS alone | Positive | GS identified a P/LP variant. TGP did not and gene of interest was analyzed. |
| *HSD17B10* (HGNC:4800) | SNV | No | GS alone | Positive | Variant classifications differed between GS and TGP reports. |
| *HUWE1* (HGNC:30892) | SNV | No | GS alone | Positive | Variant classifications differed between GS and TGP reports. |
| *IKBKG* (HGNC:5961) | Intronic | No | GS alone | Likely positive | GS identified a VUS determined to be causal. TGP did not and gene of interest was analyzed. |
| *KCNQ2* (HGNC:6296) | SNV | No | TGP alone | Likely positive | Variant classifications differed between GS and TGP reports. |
| *KCNQ3* (HGNC:6297) | SNV | No | GS alone | Positive | Variant classifications differed between GS and TGP reports. |
| *NEXMIF* (HGNC:29433) | SNV | Yes | TGP alone | Likely positive | TGP identified a P/LP variant. GS did not. |
| *SCN1A* (HGNC:10585) | SNV | No | TGP alone | Likely positive | Variant classifications differed between GS and TGP reports. |
| *SCN8A* (HGNC:10596) | Intronic | No | TGP alone | Likely positive | Variant classifications differed between GS and TGP reports. |
| *SCN8A* (HGNC:10596) | SNV | Yes | GS alone | Likely positive | GS identified a P/LP variant. TGP did not and gene of interest was analyzed. |
| *SCN8A* (HGNC:10596) | SNV | No | GS alone | Likely positive | Variant classifications differed between GS and TGP reports. |
| *SCN8A* (HGNC:10596) | SNV | No | GS alone | Positive | Variant classifications differed between GS and TGP reports. |
| *SETD5* (HGNC:25566) | SNV | No | GS alone | Positive | Variant classifications differed between GS and TGP reports. |
| *SHANK3* (HGNC:14294) | Indel | No | GS alone | Positive | GS identified a P/LP variant. TGP did not and gene of interest was analyzed. |
| *SOX5* (HGNC:11201) | CNV | No | GS alone | Positive | GS identified a P/LP variant. TGP did not and gene of interest was analyzed. |
| *SOX5* (HGNC:11201) | CNV | No | GS alone | Positive | GS identified a P/LP variant. TGP did not and gene of interest was analyzed. |
| *TGFBI* (HGNC:11771), *SOS1* (HGNC:11187) | SNV, intronic | No | GS alone | Likely positive | GS identified a VUS determined to be causal. TGP did not and gene of interest was analyzed. |
| *TSC2* (HGNC:12363) | Intronic | Yes | GS alone | Likely positive | GS identified a VUS determined to be causal. TGP did not and gene of interest was analyzed. |
| Chr7:150947163_150948277del | CNV | No | GS alone | Positive | GS identified a P/LP variant. TGP did not and gene of interest was analyzed. |
| Chr9:137661384_137714409del + ASXL3 | CNV (+ SNV) | No | GS alone | Positive | GS identified a P/LP variant. TGP did not and gene of interest was analyzed. |
| Chr15:22804175_30375696dup | CNV | Yes | GS alone | Positive | GS identified a CNV. TGP identified gene duplications or deletions and recommended high-resolution array. |
| Chr16:(38502679_46385317)_61223349dup | CNV | No | GS alone | Positive | GS identified a CNV. TGP identified gene duplications or deletions and recommended high-resolution array. |
| Chr16:(15308141_15397565)_(16367011_16403168)dup | CNV | No | TGP alone | Positive | Variant classifications differed between GS and TGP reports. |
| Chr18:1_15400036del | CNV | No | GS alone | Positive | GS identified a CNV. TGP identified gene duplications or deletions and recommended high-resolution array. |
| Chr20:63255263_63498365del | CNV | No | GS alone | Positive | GS identified a CNV. TGP identified gene duplications or deletions and recommended high-resolution array. |
| ChrX:18592341_18596203del | CNV | Yes | GS alone | Likely positive | GS identified a P/LP variant. TGP did not and gene of interest was analyzed. |

CNV, copy number variant; GS, genome sequencing; Indel, insertion/deletion; P/LP, pathogenic/likely pathogenic; SNV, single nucleotide variant; TGP, targeted gene panel

**eTable 9.** *In silico* analysis of diagnostic yield using a combination of seven commercially available targeted gene panels

| **Genetic test modality** | **NYCKidSeq study, N (%)** | ***In silico* analysis, N (%)** |
|---|---|---|
| TGP alone | 7 (1.1) | 5 (0.8)* |
| GS alone | 61 (9.5) | 41 (6.4) |
| TGP and GS | 45 (7.0) | 65 (10.1) |
| Diagnostic yield by genetic test modality | | |
| TGP yield | 52 (8.1) | 70 (10.9) |
| GS yield | 106 (16.5) | 106 (16.5) |
| **Total yield** | **113 (17.6)** | **111 (17.3)** |

* The two TGP-only variants not detected by this combination of commercially available targeted gene panels were *NEXMIF* c.2030C>A (mosaic) and Chr16:(15308141_15397565)_(16367011_16403168)dup (CNV)

| Variant type | TGP alone | TGP and GS | GS alone |
|---|---|---|---|
| CNV | 0 | 2 | 12 (2) |
| Indel | 1 | 15 | 9 |
| Intronic | 0 | 10 | 4 (1) |
| SNV | 1 (1) | 29 (1) | 23 (3) |
| Multiple types | 0 | 0 | 7 |
| | **2** | **56** | **55** |

| **Variant 1** | **Variant 2** |
|---|---|
| SNV | CNV |
| SNV | CNV |
| SNV | CNV |
| SNV | CNV |
| SNV | Intronic |
| SNV | Intronic |
| Indel | CNV |

**eFigure 1.** Concordance of detected variants between targeted gene panel (TGP) testing and genome sequencing (GS) in 113 probands who received a molecular diagnosis. The number and types of variants detected by TGP testing, GS, or both test modalities are shown. In contrast with **Figure 2**, here we considered 11 cases with variant classification differences (see **eTable 7**) as being identified by both test modalities. Eight mosaic variants were identified, including six by GS alone, one by TGP testing alone, and one by both modalities; these are indicated in parentheses for each test modality and variant type. There were seven probands for whom two distinct variants were identified (*i.e.*, multiple types). CNV, copy number variant; Indel, insertion/deletion; SNV; single nucleotide variant.
